# TB Prevalence Influence on Covid-19 Mortality

**DOI:** 10.1101/2020.05.05.20092395

**Authors:** Tareef Fadhil Raham

## Abstract

**Background:** TB latent infection reflected as TB prevalence might give heterogeneous immunity for infection in mechanism like what happen in BCG This study the first to our knowledge, addressing TB prevalence influence and possibly an important predictor for Covid-19 mortality and its findings may help to satisfy world inquiries about diversities dilemma. This study also will address disparities raised before about variances in mortalities among countries with same BCG protocols.

**Methods:** This study was set to look out for impact of TB prevalence on Covid-19 mortality on the context of countries vaccination status. Countries were divided into five groups according to BCG status. Covid-19 deaths are tested against TB prevalence through using (Non Linear Regression Modules) of predicted shapes behavior for each group.

**Results:** Slopes values have highly significant influences between TB prevalence and Covid-19 deaths for overall studied group, and among countries currently given 1 BGG (2 groups) and ones with previous history of vaccinations, being significant in currently given more than 1 BCG and in countries without vaccination. There are meaningful nonlinear regression shapes which are logarithmic in whole countries and in countries with current just 1 vaccine setting. It is inverse in other 2 groups currently given vaccine. It is power and cubic in countries never given and with previously given vaccines respectively. All groups and whole sample shows either perfect or extremely perfect R-square (Determination Coefficient) values with significant in at least at P-values<0.05. Study denotes possibility of factor/s other than BCG prevalence (i.e. The intercept) were operating in different ranges within groups.

**Conclusion:** High TB prevalence together with continuing BCG programs decrease COVID-19 Mortalities in different countries.

**Strengths and limitations of this study:**

- To our knowledge, this study will be the first addressing TB Prevalence influence and possibly an important predictor for Covid-19 mortality and its findings may help to satisfy world inquiries about diversities dilemma.
- This study also will address disparities raised before about variances in mortalities among countries with same BCG protocols.
- Potential confounding factors still exist.

## Introduction

*A*bout one-quarter of the world’s population may be infected with latent tuberculosis^1^ which is defined as a state of persistent immune response to stimulation by Mycobacterium tuberculosis antigens with no evidence of clinically manifest active TB^2^. It is estimated that the lifetime risk of an individual with LTBI for progression to active TB is 5–10%^3^, furthermore more than 90% of people infected with *M. tuberculosis* for more than two years never develop tuberculosis even if their immune system is severely suppressed^4^, while BCG is highly efficacious at preventing meningeal and miliary TB, but is at best 60% effective against the development of pulmonary TB in adults and wanes with age.^5^ It is hypothesized that beneficial heterologous non-specific effects and innate immune memory training through epigenetic reprogramming.^5^,^6^ Furthermore secretion of pro-inflammatory cytokines, specifically IL-1B play a vital role in antiviral immunity this is believed to be increased by intradermal inoculation of live attenuated Mycobacterium bovis through BCG vaccine.^7^ Currently, two clinical trials are ongoing to determine if BCG vaccination protects healthcare workers during the covid-19 pandemic^8^. Review by WHO’s Strategic Advisory Group of Experts on Immunization (SAGE) on non-specific effects of BCG vaccine shows that vaccination at birth reduces neonatal mortality by 48% (18–67%), which is mainly due to the prevention of neonatal sepsis and respiratory infections^9^. Variances in Covid-19 incidence and mortality across the world led people to look for possible cause (s). One thing (among others) is BCG vaccine as far as being subjected to different protocols and policies among different nations. Rapid systematic review and WHO statements regarding vaccine protocols concluded that current evidence does not justify changes to current global immunization policy and they recommend further studies of adequate size and quality to determine the non-specific effects of vaccines on all-cause mortality and findings that show significant correlation between BCG vaccine and Covid-19 provide inadequate evidence and should not be reflected on any practices or policies at the current time, outside the contexts of RCTs.^8^

These recommendations did not handle the current latent TB protocols variances among nations. Based on the underlying hypotheses of BCG vaccine, I try to look for this from different view which is the beneficial effect of mycobacterium infection through latent infection which carry a natural BCG like effect to the host and reflected indirectly by the prevalence of TB in the community taking in consideration more beneficial effects of latent infection over BCG vaccination as mentioned earlier and I put the question whether variances in prevalence is reflected on Covid-19 mortality or not in order to support or reject the BCG and TB prevalence hypotheses.

## Material and methods

### Data regarding

TB prevalence were taken from publically published WHO data while Covid-19 data 2018 were taken from publically published data collected between 30/4-2/5/2020. Every country has less than 1 million populations is excluded also Lebanon is excluded due to non-representative sample due to refugees and immigration to and out Lebanon.

Total number of groups is 155 Country according to BCG criteria shown in table No. (1).

### Patient and public involvement statement

It was not appropriate or possible to involve patients or the public in this work given that we used general practice level summated data and related publically published morbidity and mortality statistics.

Patients were not involved

## Statistical Methods

The optimum model fit quality were checked among the assumed models of Non-Linear regression, such as (Logarithmic, Inverse, Quadratic, Cubic, Compound, Power, S-Shape, Growth, Exponential, and Logistic) as well as the simple linear regression model for predicted equations with their estimates such that (Slope, Intercept, Correlation Coefficient, and regression ANOVA) are suggested for studying influence of TB Prevalence / 100000 in relative to Covid -19 mortality/million and as illustrated in the following table (2). All statistical operations were performed using the ready-made statistical package SPSS, ver. 22.

**Table (1):**
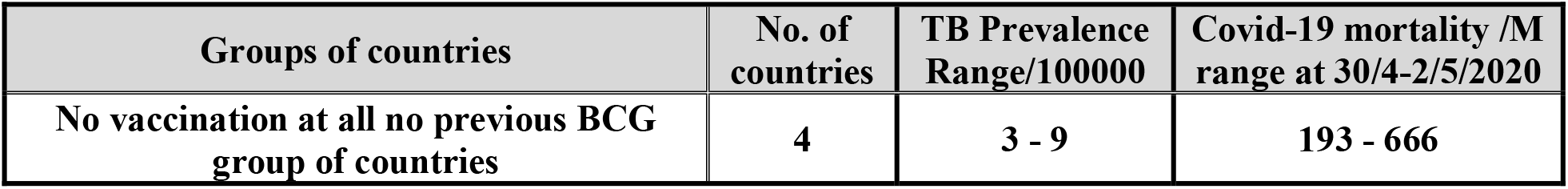
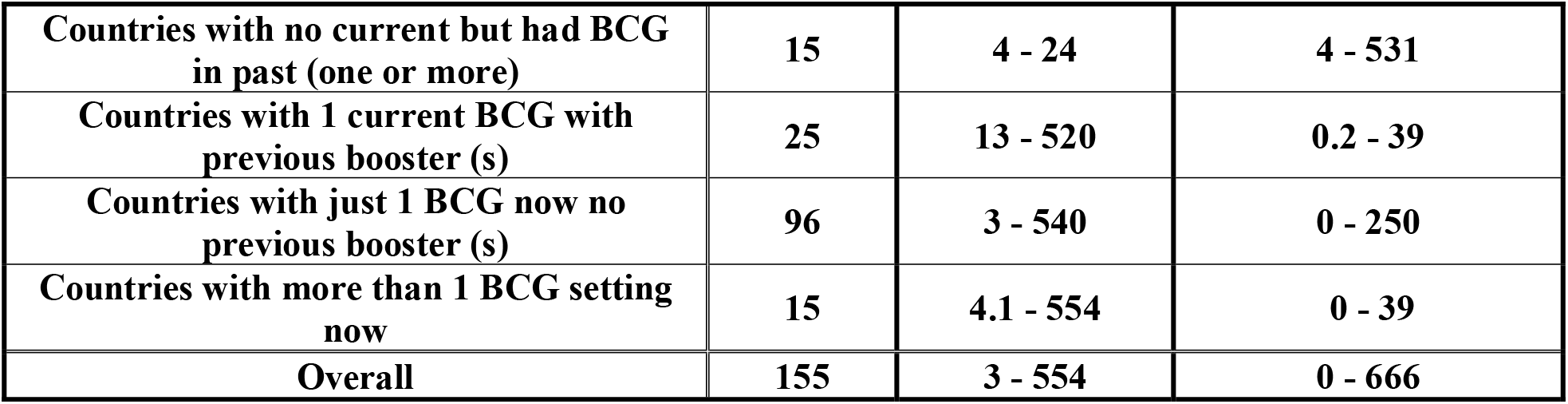
shows classification of studied groups with ranges of TB Prevalence and Covid-19 mortality.

**Table (2):**
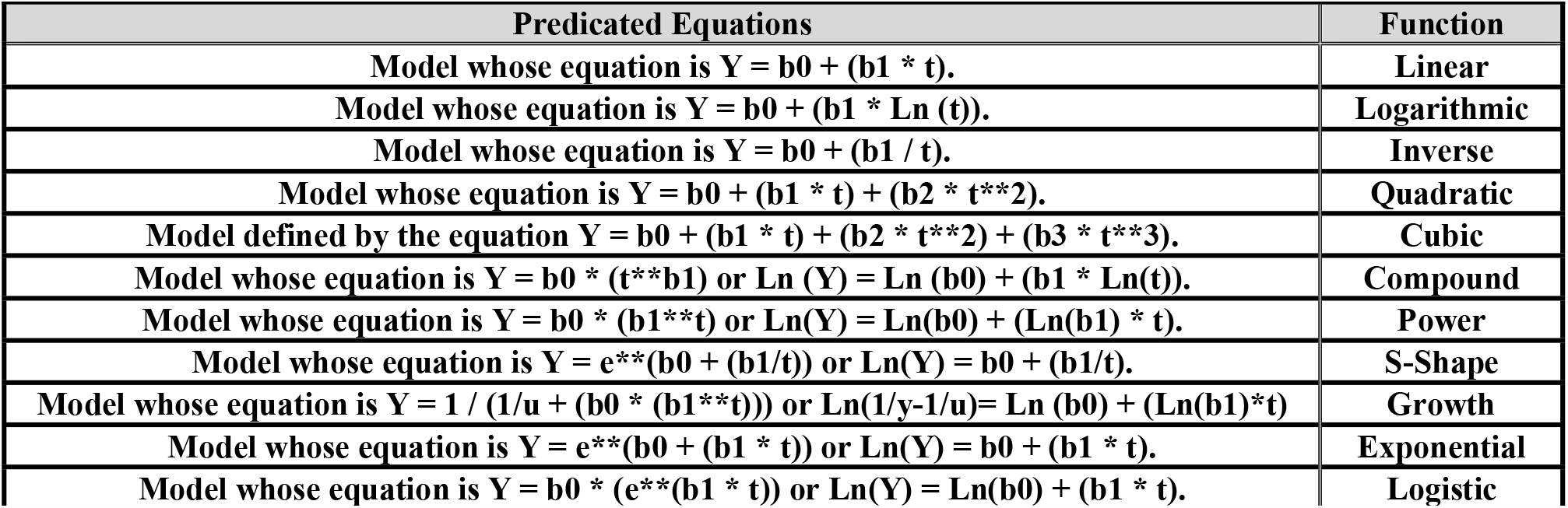
Suggested Predicated Equations for studying the Influence of TB prevalence / 100000 in relative to Covid -19 mortality / million

## Results

### 1- Influence of TB prevalence/100000 on Covid-19 mortality/million on (no vaccination at all no previous BCG) group of countries

Table (3) and figure (1) show meaningful nonlinear regression of (power model) tested in two tailed alternative statistical hypotheses. Slope value indicates that with increasing one unit of the first factor, there is positive influence on unit of the second factor, and estimated by (1.128). The increment recorded significant influence at P<0.05, as well as relationship coefficient at P<0.05 are accounted (0.987) with extremely perfect meaningful determination coefficient (R-Square = 97.45%). Other source of variations that are not included in model, i.e. “Constant term in regression equation” shows that non assignable factors that not included in the regression equation, ought to be informative since 52 cases are in expectation without the effect of the factors tested (with significant p value) which indicates valuable other factor (s) not included in model as shown in table (3). Figure (1) also shows the long term trend of scatter diagram influence of TB prevalence related to Covid -19 mortality for the group.

**Table (3):**
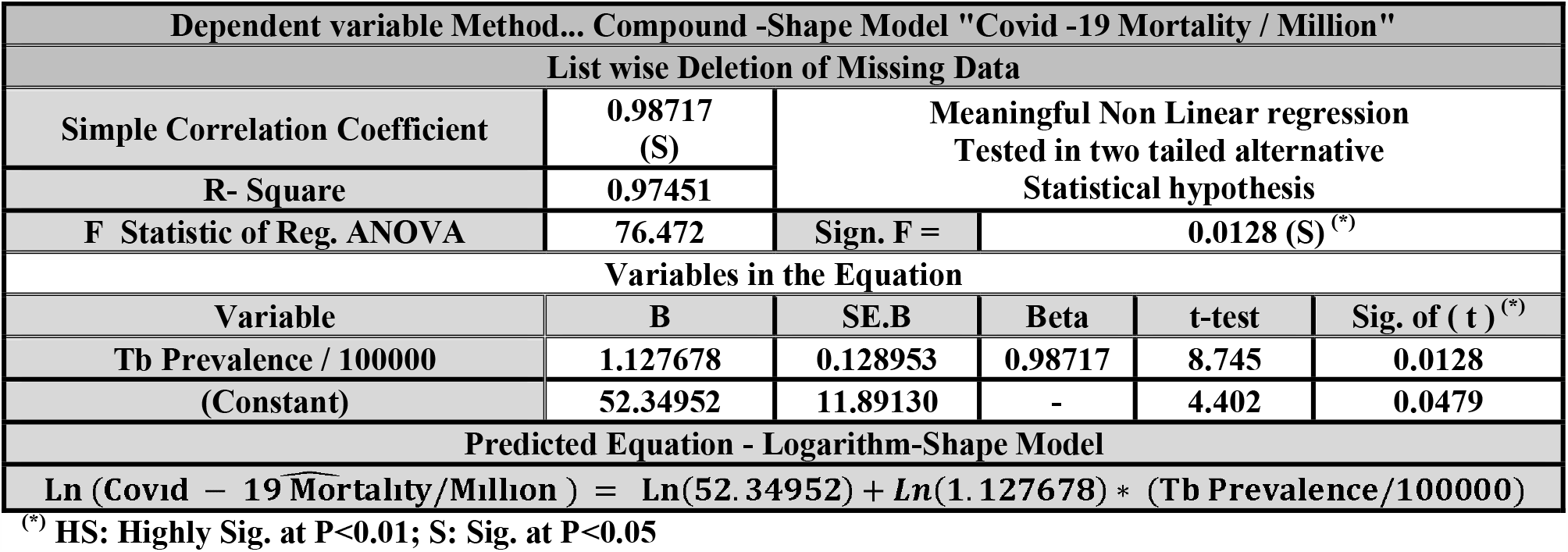
Influence of TB prevalence / 100000 factor on Covid -19 mortality / million factor for (no vaccination at all no previous BCG) group

**Figure (1):**
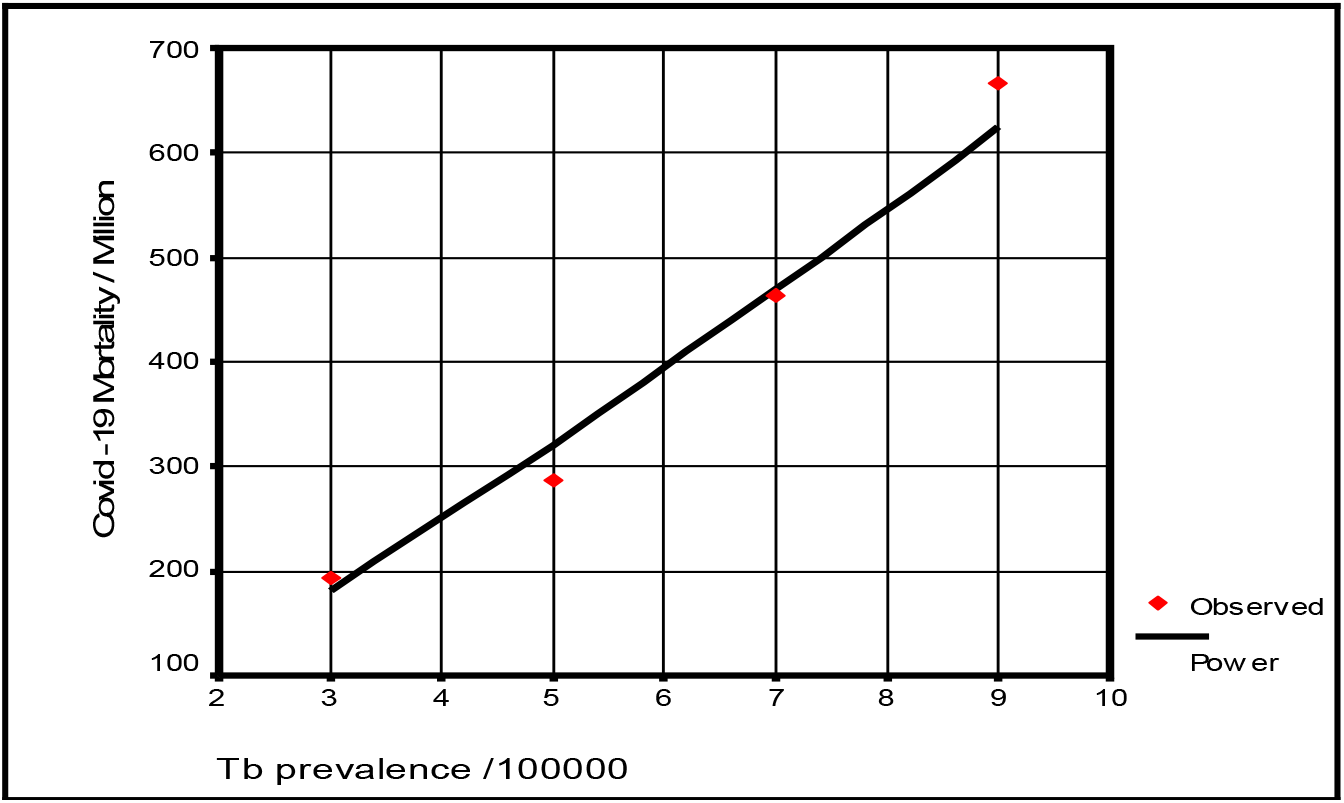
Long term trend of scatter diagram influence concerning TB prevalence /100000 with Covid-19 mortality/million for no vaccination at all no previous BCG group

### 2- Influence of TB prevalence/100000 on Covid - 19 mortality/million on (no current but had BCG in past (one or more)) group of countries

Table (4) figure (2) shows meaningful nonlinear regression of (Cubic model) tested in two tailed alternative statistical hypotheses. Slopes values are estimated and indicates that with increasing one unit of the first factor, there is high increments with the second factor up to marginal of 9 TB prevalence rate influence and then decreasing behavior with leftover of the second factor, recording high significant influence at P<0.01, as well as relationship coefficient accounted (0.81301) at P<0.01 with extremely perfect meaningful determination coefficient (R-Square = 66.098%). Others source of variations that are not included in model, i.e. “Constant term in regression equation” shows that non assignable factors that are not included in the regression equation, ought to be informative, with a high significance at P<0.01. Figure (2) also shows the long term trend of scatter diagram influence of TB prevalence in relation to Covid - 19 million for the group.

**Table (4):** Influence of TB prevalence / 100000 factor on Covid - 19 mortality / million factor for (no current but had BCG in past or more) group

| Dependent variable Method... Polynomial Cubic -Shape Model "Covid -19 Mortality / Million" |  |  |  |  |  |
| --- | --- | --- | --- | --- | --- |
| List wise Deletion of Missing Data |  |  |  |  |  |
| Simple Correlation Coefficient | 0.81301 (HS) | Meaningful Non Linear regression<br>Tested in two tailed alternative Statistical hypothesis |  |  |  |
| R- Square | 0.66098 |  |  |  |  |
| F Statistic of Reg. ANOVA | 11.698 | Sign. F = | 0.0015 (HS) <sup>(*)</sup> |  |  |
| Variables in the Equation |  |  |  |  |  |
| Variable | B | SE.B | Beta | t-test | Sig. of ( t ) <sup>(*)</sup> |
| Tb Prevalence / 100000 | 107.3544 | 22.25379 | 3.21713 | 4.824 | 0.0004 |
| Tb Prevalence / 100000^3 | -0.144688 | 0.030415 | -3.1727 | -4.757 | 0.0005 |
| (Constant) | -474.92341 | 135.603 | - | -3.502 | 0.0044 |
| Predicted Equation - Logarithm-Shape Model |  |  |  |  |  |
| (Covid – 19 Mortality/Milhon)<br>= (−474. 92) + (107. 354) * (Tb Prevalence/100000) − (0. 144688) * (Tb Prevalence/100000)^3 |  |  |  |  |  |

<sup>(\*)</sup> HS: Highly Sig. at $P < 0.01$ ; S: Sig. at $P < 0.05$

**Figure (2):**
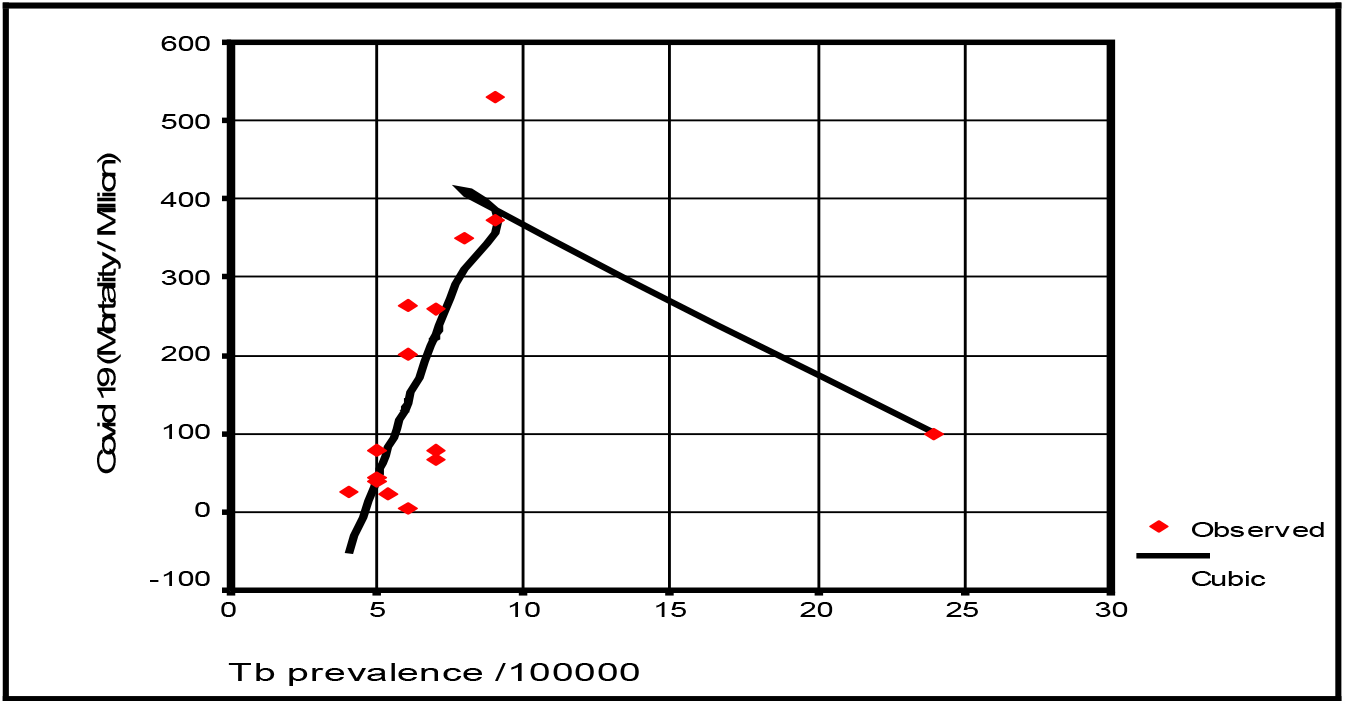
Long term trend of scatter diagram influence concerning TB prevalence /100000 with Covid-19 mortality/million for no current but had BCG in past or more group

### 3- Influence of TB prevalence/100000 on Covid - 19 mortality/million on 1 current BCG and previous booster (s) group of countries

Table (5) and (figure 3) show meaningful nonlinear regression of (inverse model) tested in two tailed alternative statistical hypothesis of two factors. Slope value indicates increasing one unit of the first factor, there is a reverse influence on the unit of the second factor, and estimated as (465), and that recorded increment is highly significant at P<0.01, as well as relationship coefficient at P<0.01, and accounted (0.63662) with meaningful determination coefficient (R-Square). Other source of variations that are not included in model, i.e. “Constant term in regression equation” shows that non assignable factors that are not included in the regression equation, could be neglected, since estimated (-1.202066) mortality expected initially without effectiveness by the group with no significant at P>0.05.

**Table (5):** Influence of TB prevalence / 100000 factor on Covid - 19 mortality / Million factor of 1 current BCG + previous booster (s)

| Dependent variable Method... Inverse-Shape Model "Covid -19 Mortality / Million" |  |  |  |  |  |
| --- | --- | --- | --- | --- | --- |
| List wise Deletion of Missing Data |  |  |  |  |  |
| Simple Correlation Coefficient | 0.63662 (HS) | Meaningful Non Linear regression Tested in two tailed alternative Statistical hypothesis |  |  |  |
| R- Square | 0.40529 |  |  |  |  |
| F Statistic of Reg. ANOVA | 14.08977 | Sign. F = | 0.0008 (HS) <sup>(*)</sup> |  |  |
| Variables in the Equation |  |  |  |  |  |
| Variable | B | SE.B | Beta | t-test | Sig. of ( t ) <sup>(*)</sup> |
| Tb Prevalence / 100000 | 465.0560 | 117.7817 | 0.636625 | 3.872 | 0.0008 |
| (Constant) | -1.202066 | 4.830430 | - | -0.249 | 0.8058 |
| Predicted Equation - Logarithm-Shape Model |  |  |  |  |  |
| Covid - 19 Mortality / Million = $-1.2021 - [465.0560 / (Tb\ Prevalence / 100000)]$ | | | | | |

<sup>(\*)</sup> HS: Highly Sig. at $P<0.01$ ; S: Sig. at $P<0.05$

**Figure (3):**
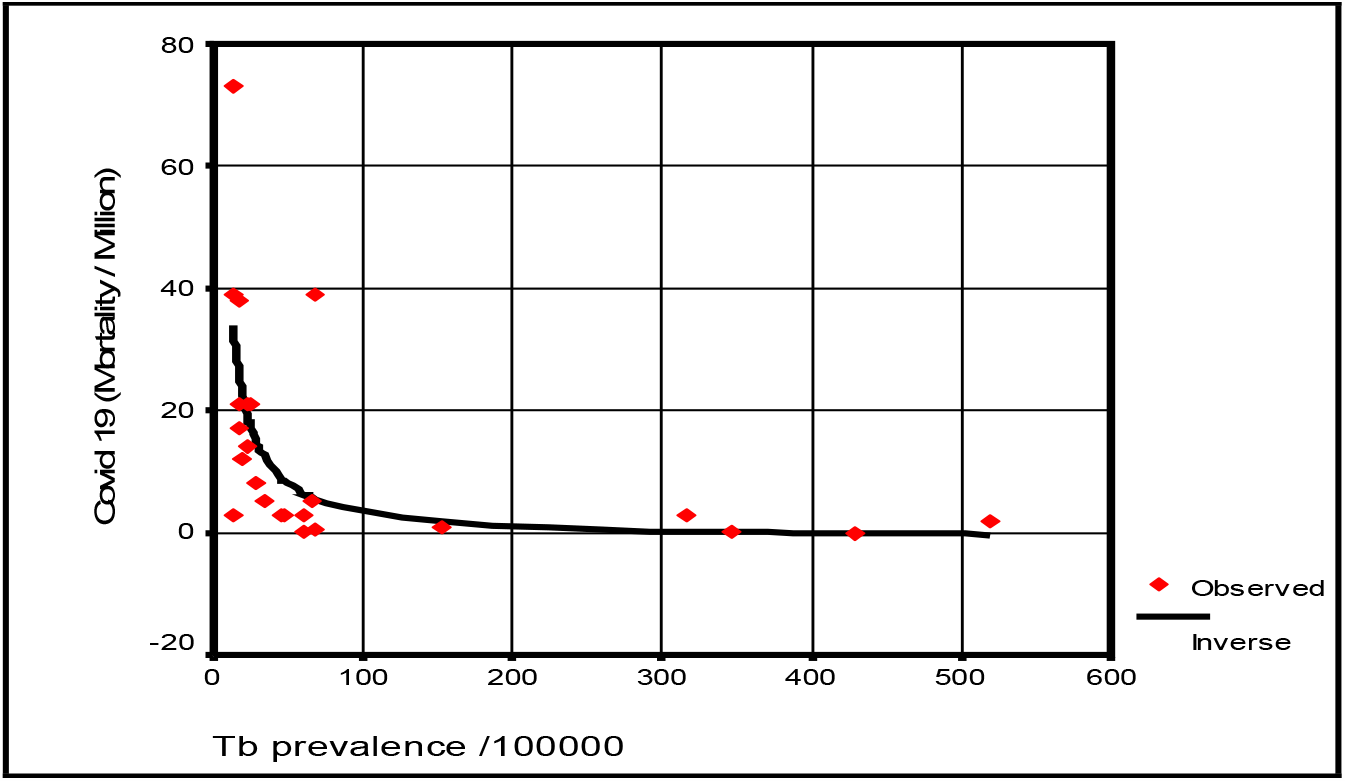
Long term trend of scatter diagram influence concerning TB prevalence /100000 with Covid 19 mortality/million for 1 current BCG with previous booster (s) group

Figure (3) also shows the long term trend of scatter diagram influence of TB prevalence in relation to Covid - 19 mortality for the group.

### 4- Influence of TB prevalence/100000 on Covid - 19 mortality/million on (just 1 BCG now no previous booster (s)) group

Table (6) shows meaningful nonlinear regression of the group factors (Logarithm model) as shown in figure (4). Slope value indicates that with increasing one unit of the first factor, occurrence negative influence on unit of the second factor, and estimated by (-2.513626), and this is highly significant with reverse influence at P<0.01, as well as relationship coefficient at P<0.01 is recorded between studied factors, and counted as (0.30751) with meaningful determination coefficient (R-Square). Others source of variations not included in model, i.e. “Constant term in regression equation” shows that non assignable factors that not included in the regression equation, ought to be informative with highly significant at P<0.01as shown in table (6).

**Table (6):** Influence of TB prevalence / 100000 factor on Covid - 19 mortality / million factor of just 1 BCG now no previous booster (s) group

| Dependent variable Method... Logarithm-Shape Model "Covid -19 Mortality / Million" |  |  |  |  |  |
| --- | --- | --- | --- | --- | --- |
| List wise Deletion of Missing Data |  |  |  |  |  |
| Simple Correlation Coefficient | 0.30751 (HS) | Meaningful Non Linear regression<br>Tested in two tailed alternative<br>Statistical hypothesis |  |  |  |
| R- Square | 0.09456 |  |  |  |  |
| F Statistic of Reg. ANOVA | 9.81702 | Sign. F = | 0.0023 (HS) <sup>(*)</sup> |  |  |
| Variables in the Equation |  |  |  |  |  |
| Variable | B | SE.B | Beta | t-test | Sig. of ( t ) <sup>(*)</sup> |
| Tb Prevalence / 100000 | -2.513626 | 0.802252 | -0.30751 | -3.133 | 0.0023 |
| (Constant) | 15.458213 | 3.534547 | - | 4.373 | 0.0000 |
| Predicted Equation - Logarithm-Shape Model |  |  |  |  |  |
| Covid – 19 Mortality / Million = 15.458 – 2.514 Ln(Tb Prevalence/100000) |  |  |  |  |  |

<sup>(\*)</sup> HS: Highly Sig. at $P < 0.01$ ; S: Sig. at $P < 0.05$

**Figure (4):**
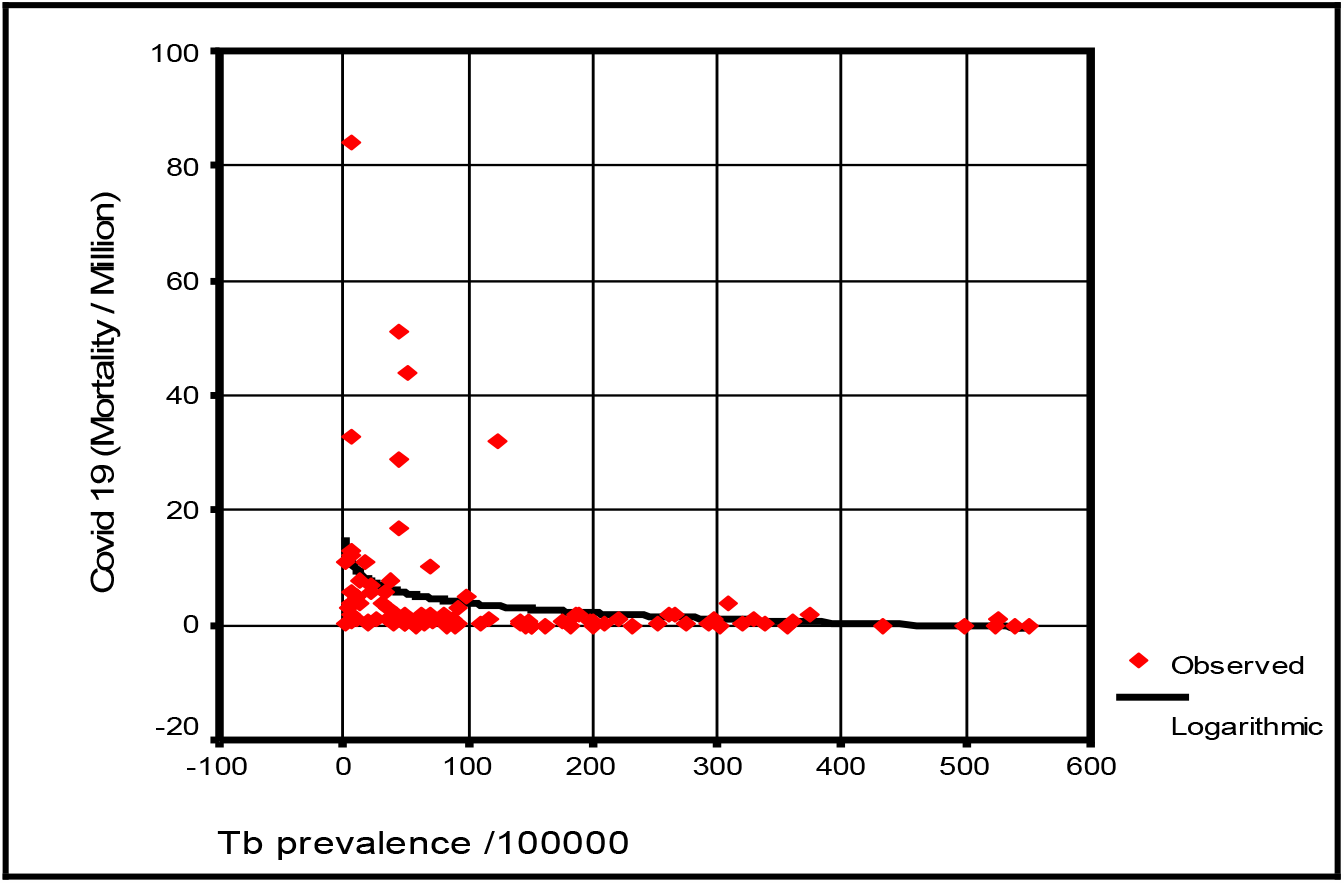
Long term trend of scatter diagram influence concerning TB prevalence with Covide-19 mortality for (just 1 BCG now, no previous booster (s)) group

Figure (4) also shows the long term trend of scatter diagram influence of TB prevalence in relation to Covid - 19 mortality for the group.

### 5- Influence of TB Prevalence/100000 on Covid - 19 Mortality/Million on more than (1 BCG setting now) group countries

Table (7) and figure (5) show meaningful nonlinear regression of (Inverse model) tested in two tailed alternative statistical hypothesis of two factors. Slope value indicates that with increase one unit of the first factor, there is a reverse influence on the unit of the second factor, and estimated by (138), and that increment recorded significant at P<0.05, as well as relationship coefficient at P<0.05 are recorded between studied factors, and is accounted (0.61580) with meaningful determination coefficient (i.e. R-Square). Other source of variations are not included in model, i.e. “Constant term in regression equation” shows that non assignable factors that not included in the regression equation, could be neglected, since estimated (7.555379) cases of Covid - 19 mortality / million expected initially without effectiveness by the group more than 1 BCG setting now, with no significant at P>0.05.

**Table (7):** Influence of Tb Prevalence / 100000 factor on Covid - 19 Mortality / Million factor of more than 1 BCG setting now

| Dependent variable Method... Inverse-Shape Model "Covid -19 Mortality / Million" |  |  |  |  |  |
| --- | --- | --- | --- | --- | --- |
| List wise Deletion of Missing Data |  |  |  |  |  |
| Simple Correlation Coefficient | 0.61580 (S) | Meaningful Non Linear regression Tested in two tailed alternative Statistical hypothesis |  |  |  |
| R- Square | 0.33146 |  |  |  |  |
| F Statistic of Reg. ANOVA | 7.94114 | Sign. F = | 0.0145 (S) <sup>(*)</sup> |  |  |
| Variables in the Equation |  |  |  |  |  |
| Variable | B | SE.B | Beta | t-test | Sig. of ( t ) <sup>(*)</sup> |
| Tb Prevalence / 100000 | 137.73131 | 48.8755 | 0.615802 | 2.818 | 0.0145 |
| (Constant) | 7.555379 | 3.66279 | - | 2.063 | 0.0597 |
| Predicted Equation - Logarithm-Shape Model |  |  |  |  |  |
| Covid – 19 Mortality / Milhon = 7.555379 – [137.73131 / (Tb Prevalence/100000)] |  |  |  |  |  |

(\*) HS: Highly Sig. at $P<0.01$ ; S: Sig. at $P<0.05$ ; NS: Non Sig. at $P>0.05$

**Figure (5):**
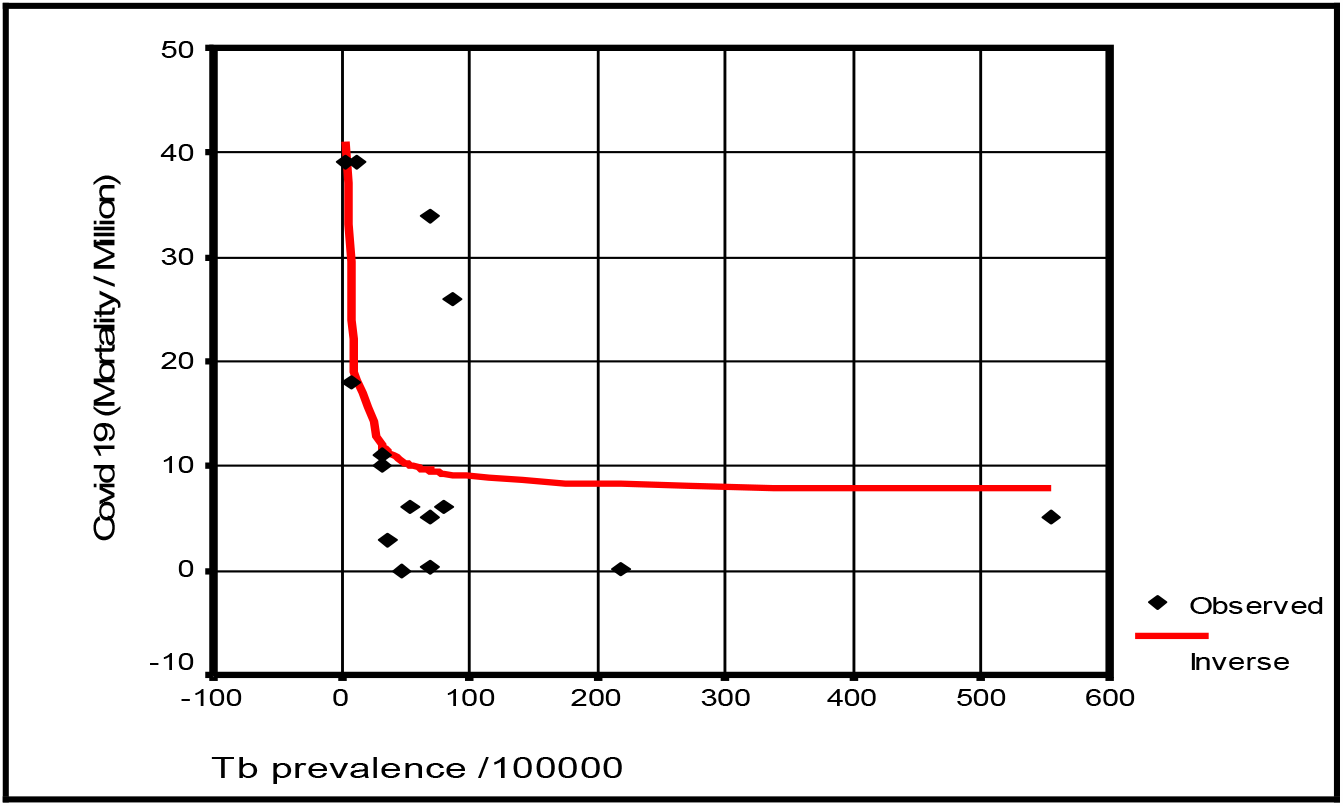
Long term trend of scatter diagram influence concerning Tb prevalence /100000 with Covide 19 mortality/million of more than 1 BCG setting now

Figure 5 also shows the long term trend of scatter diagram influence of Tb prevalence in relation to Covid - 19 mortality for the group.

### 6- Influence of TB Prevalence/100000 on Covid-19 mortality/million on overall countries

Table (8) and figure (6) show meaningful nonlinear regression of (Logarithm model) tested in two tailed alternative statistical hypothesis of two factors tested on all countries (not restricted to any group). Slope value indicates that with increasing one unit of the first factor, there is negative influence on unit of the second factor, and estimated by (-23.49066), and that increment recorded as highly significant with reverse influence at P<0.01, as well as highly significant relationship coefficient at P<0.01 is recorded between studied factors, and accounted (0.36749) with meaningful determination coefficient (R-Square). Other source of variations not included in studied model, i.e. “Constant term in regression equation” shows that non assignable factors that are not included in the regression equation, ought to be informative with highly significant P value <0.01, which indicates that rather than meaningful and significant relationship between studied factors, a meaningful and high significant effectiveness of others factors that are not included in the predicted model on occurrences of Covid - 19 mortality.

**Table (8):**
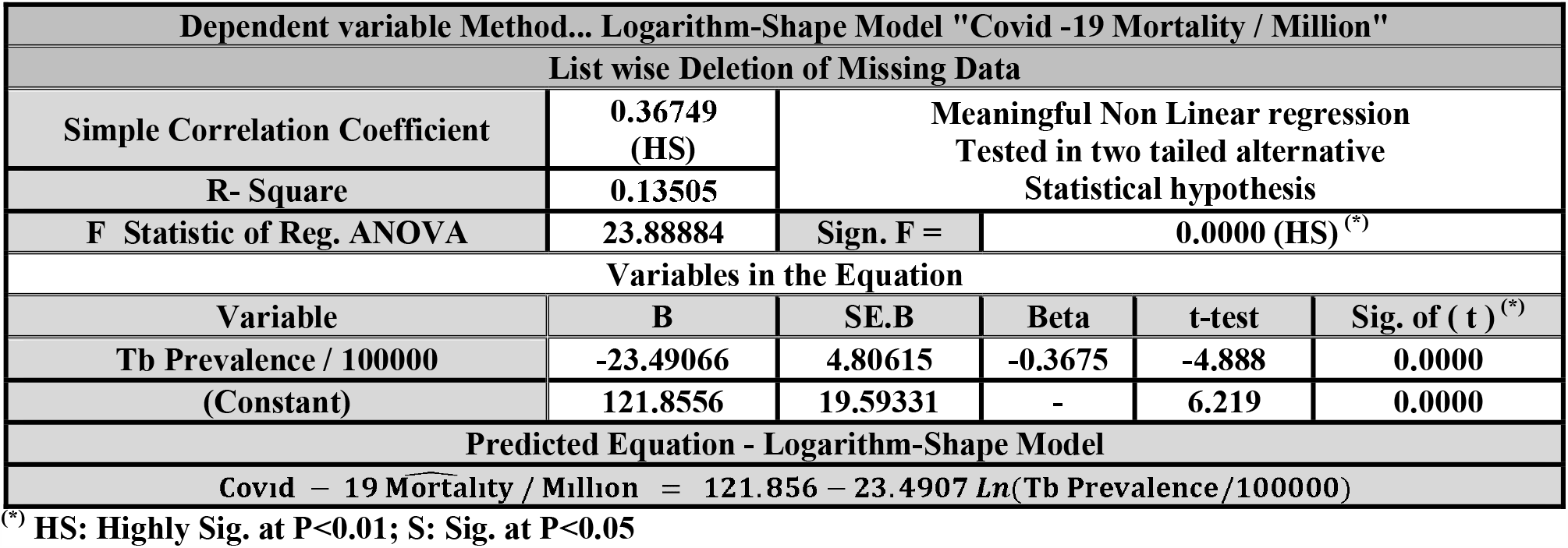
Influence of TB Prevalence / 100000 factor on Covid-19 mortality / million factor of an overall groups

| Dependent variable Method... Logarithm-Shape Model "Covid -19 Mortality / Million" |  |  |  |  |  |
| --- | --- | --- | --- | --- | --- |
| List wise Deletion of Missing Data |  |  |  |  |  |
| Simple Correlation Coefficient | 0.36749<br>(HS) | Meaningful Non Linear regression<br>Tested in two tailed alternative<br>Statistical hypothesis |  |  |  |
| R- Square | 0.13505 |  |  |  |  |
| F Statistic of Reg. ANOVA | 23.88884 | Sign. F = | 0.0000 (HS) <sup>(*)</sup> |  |  |
| Variables in the Equation |  |  |  |  |  |
| Variable | B | SE.B | Beta | t-test | Sig. of ( t ) <sup>(*)</sup> |
| Tb Prevalence / 100000 | -23.49066 | 4.80615 | -0.3675 | -4.888 | 0.0000 |
| (Constant) | 121.8556 | 19.59331 | - | 6.219 | 0.0000 |
| Predicted Equation - Logarithm-Shape Model |  |  |  |  |  |
| Covid – 19 Mortality / Million = 121.856 – 23.4907 Ln(Tb Prevalence/100000) |  |  |  |  |  |

<sup>(\*)</sup> HS: Highly Sig. at P<0.01; S: Sig. at P<0.05

**Figure (6):**
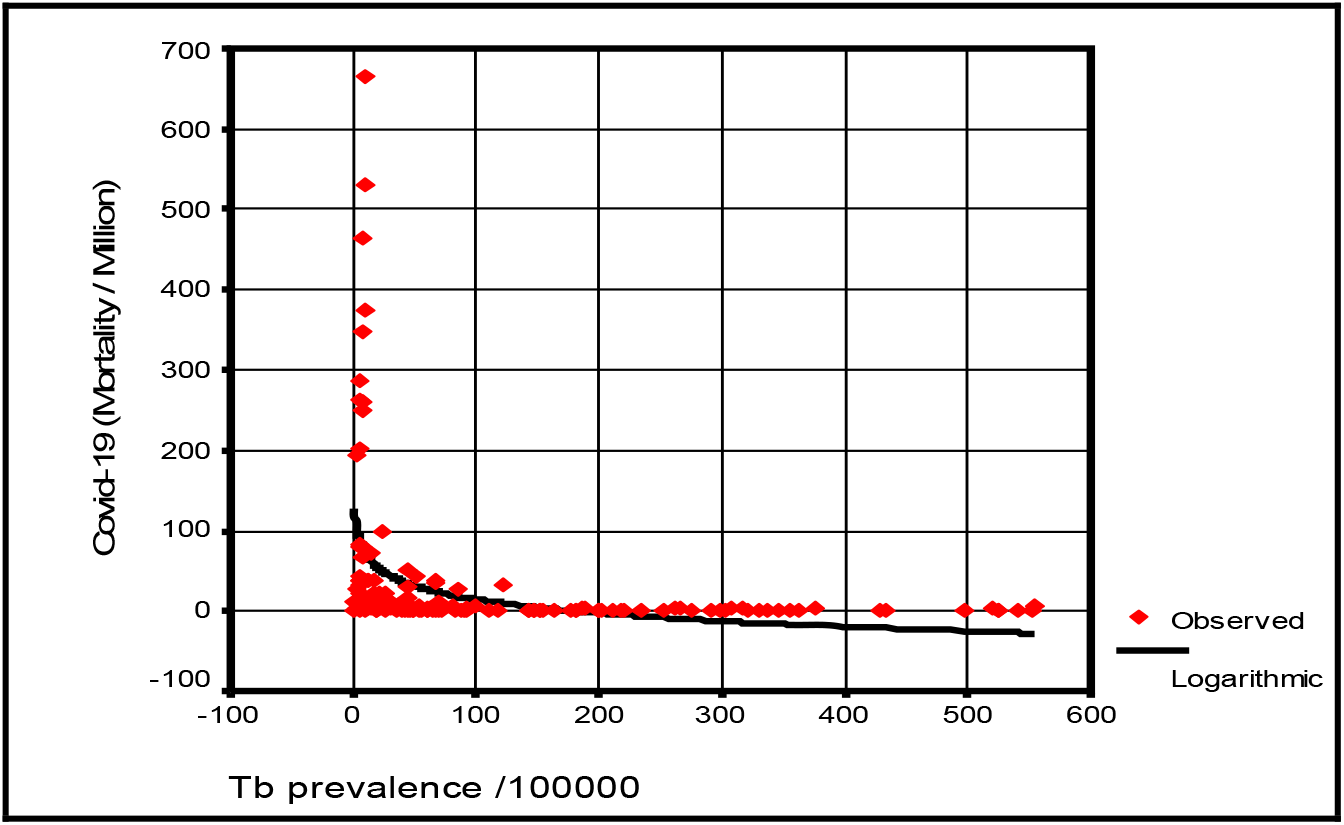
Long term trend of scatter diagram influence of TB prevalence /100000 with Covid-19 mortality/million of an overall groups.

Figure (6) also show the long term trend of scatter diagram influence of TB prevalence in whole countries tested.

## Discussion

Lebanon about 6.830 million was excluded due recent 1.5 million refugees from Syria and 400,000 Palestinian refugees in addition to 1 million foreign workers (mainly Syrians) and there is a sizeable Lebanese diaspora. Countries with just thousands of people up to less than 1 million populations are excluded because it is impossible to look for 1 million mortalities for Covid 19 unless adjusting figures by augmenting population figures.

Our data suggest that TB prevalence seems to significantly reversely associated with reduction in mortality associated with Covide-19 this finding is enhanced by BCG status being the more frequent the vaccine given the less Covid-19mortality is. As far as active TB infection reflects 10 % iceberg of total TB infection, latent TB possibly works with same mechanism that BCG works in provocation immunity towards viruses. Strong association in this study might reflect a more potent effect of prevalence over BCG.

The strong negative association between Covid -19 mortality and TB prevalence in many tables is going parallel with underling hypotheses of our study and furtherly support BCG theory which was tested many weeks before.

Significant inverse and logarithmic associations among tablets reflect augmentation effect of vaccination and possibly effect of revaccination these finding support the previously mentioned hypotheses by different way and may raise the question of early interventions before clinical control trials result proceed, specially these trials will take a time while urgent actions are needed to contain the disease in context of available evidence at this time. The possible actions according to this study include questioning BCG practices as previous studies recommends and extends to policies whether to treat latent TB infections or not specially the WHO recommends tailored latent tuberculosis infection management based on tuberculosis burden and resource availability^10^. These make these recommendations wider than previous BCG studies in this aspect although it is based on the same underling hypotheses.

Treatment of latent TB to prevent about 5 to 10 percent of infected people who will develop TB disease if not treated is a common practice in developed countries ^11^ and this should be reevaluated.

Tuberculosis Prevalence is defined as the number of TB cases (all forms) at a given point in time^12^. and an algorithm for excluding active tuberculosis was specified in the policies of 43 (63.2%) low-burden countries^13^. The content of that algorithm varied greatly from country to country. In Ecuador the recommendation was only that active tuberculosis should be ruled out, with no mention of an exclusion algorithm.^13^ Furthermore recorded HIV prevalence is highest in Ecuador at 35% this might explains the high prevalence rate of TB and non proportioned Covid-19 mortality due to possible inclusion of latent Tb in prevalence of TB due to high HIV in absence of inclusion criteria regarding absent of exclusion algorithm.

In 2014, the global burden of LTBI was 23.0%, amounting to approximately 1.7 billion people. WHO South-East Asia, Western-Pacific, and Africa regions had the highest prevalence and accounted for around 80% of those with LTBI^14^. On the country level, China and India had the highest LTBI burden, approximately 350 million infections, followed by Indonesia at around 120 million infections and fewer than 60 million infections in all other countries. The USA had the 20th burden, at an estimated 13 million.^14^ the diversity of spread and mortality goes parallel with low latent TB places and supported by low prevalence of TB according to this study. Other confounding factors which miht play a role as study shows should be clarified in future studies.

## Conclusion

TB prevalence is strongly negatively associated with covid-19 mortality among countries and Covid-19 mortality is more sever in BCG vaccination status of whole country.

## Recommendations

Randomized clinical trials are recommended on this aspect.

Early interventions might be considered based on the supportive evidence at this time which include BCG vaccination protocols and review of current latent TB programs.

## Ethics and dissemination

Ethical permission is not necessary as this study analyzed publically published data and patients were not involved.

There is no conflict of interest.

There is no funding received.

## Data Availability

All data referred to in the manuscript are available on requist

## Acknowledgment

I am deeply grateful to Emeritus Professor Abdulkhaleq Abduljabbar Ali Ghalib Al-Naqeeb, Ph.D. in the Philosophy of Statistical Sciences at the Medical & Health Technology college, Baghdad-Iraq, for his assistance and supported in data analysis, interpretations of finding results, and revise and display the paper.

